# Left Ventricular Assist Device Outcomes and Strategy 5 Years from Heart Transplant Allocation Score Change

**DOI:** 10.1101/2023.09.11.23295392

**Authors:** Jacob Agronin, Meredith Brown, Hannah Calvelli, Val Rakita, Yoshiya Toyoda, Mohammed Abul Kashem

## Abstract

**Background:** The United Network for Organ Sharing (UNOS) adopted new criteria for the heart allocation score on 10/18/2018 to reflect changing trends of candidates’ mortality while awaiting transplant. We examined the impact of these policy changes on rates of left ventricular assist device (LVAD) implantation and outcomes posttransplant from a relatively newer UNOS database.

**Methods:** The UNOS registry was used to identify first-time adult heart recipients with LVAD at listing or transplant who underwent transplantation between 1/1/2016 and 3/10/2020. Survival data was collected through 3/30/2023. Those listed prior to 10/18/2018 but transplanted after were excluded. Patients were divided into before or after change groups. Demographics and clinical parameters were compared. Survival was analyzed with Kaplan-Meier curves and log-rank tests. A p<0.05 was considered significant.

**Results:** We identified 4599 heart recipients with LVAD in the before (N=3767) and after (N=832) score change eras. The after group had a lower rate of LVAD implantation while listed compared to the before group (19.4% vs 34.5%, p<0.0001), younger average age (53.1 ± 12.2 vs 54.1 ± 11.9, p=0.0350) and more likely to be female (24.9% vs 19.6%, p=0.0007); in both groups, most recipients (62%) were white. There was significantly farther distance from the donor hospital to transplant center in the after group (259.5 ± 246.8 NM vs 143.2 ± 182.1 NM, p<0.0001) and decreased waitlist days (83.5 ± 103.5 vs 369.0 ± 458.5, p<0.0001). Recipients in the after group were more likely to receive a CDC increased-risk donor organ (37.5% vs 30.2%, p=0.0002). Survival at 5-years was significantly reduced in the after group (60.5% vs 78.9%, p<0.0001).

**Conclusions:** The allocation score change in 2018 yielded considerable changes in mechanical circulatory support device implantation strategy and survival. The rate of LVAD implantation decreased with profoundly worse 5-year survival, showing further divergence from prior short-term post-transplant data.

## Introduction

### Background/rationale

Heart transplant allocation scoring criteria are heavily based on chance of mortality while waiting for a transplant. As mechanical circulatory support devices (MCSDs) continue to improve in terms of durability, reduced adverse events, and overall effect on survival, there was a need to rethink the prioritization of donor hearts. The United Network for Organ Sharing (UNOS) adopted new criteria for the heart allocation score in October 2018 in part to reflect changing mortality trends of heart transplant candidates implanted with MCSDs (Table 1). The scoring system changed in several ways, but notably by further stratifying the existing three categories into six and deprioritizing stable candidates with left ventricular assist devices (LVADs).^1,2^ We examined the impact of these policy changes on rates of LVAD implantation and post-transplant outcomes.

**Table 1.**
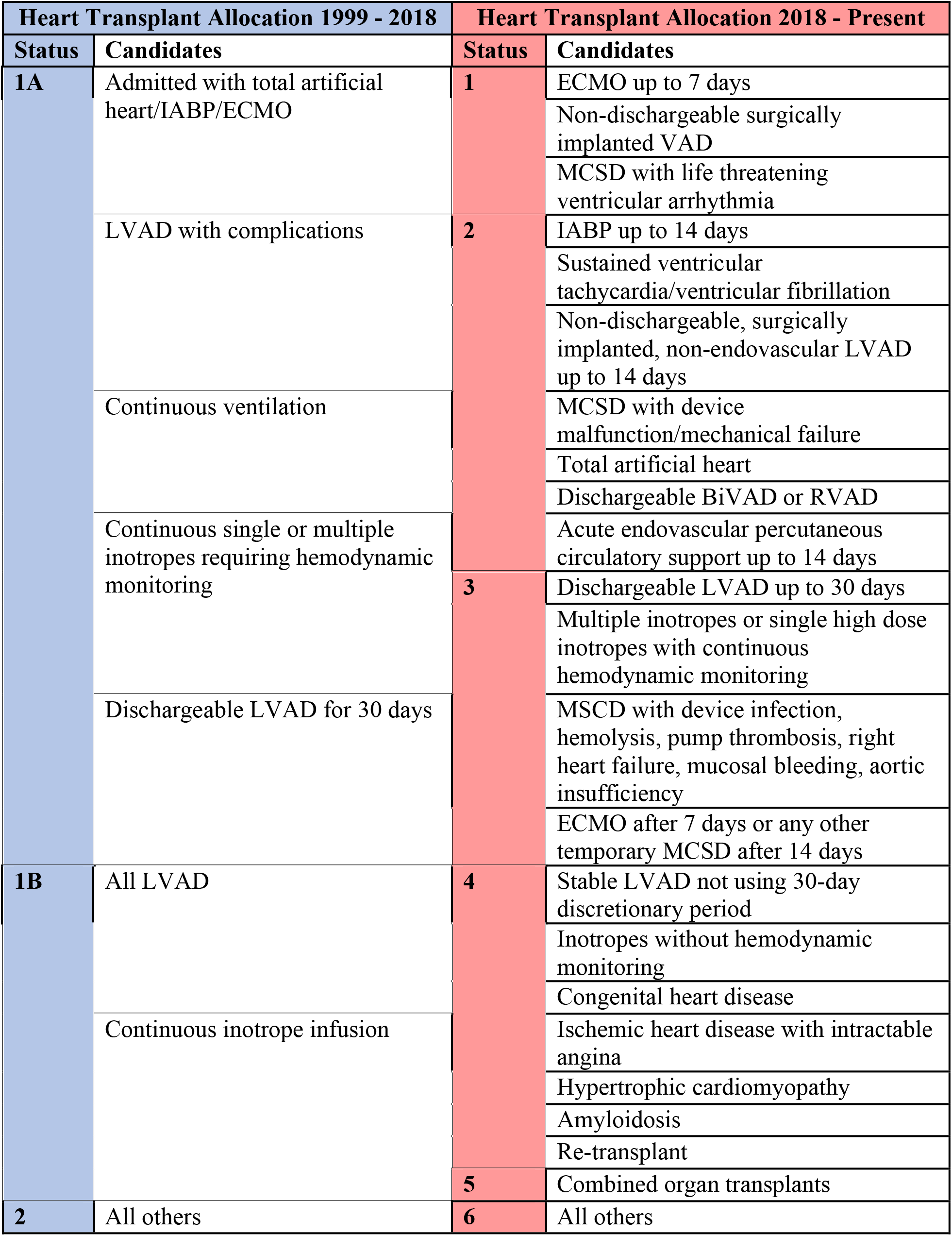
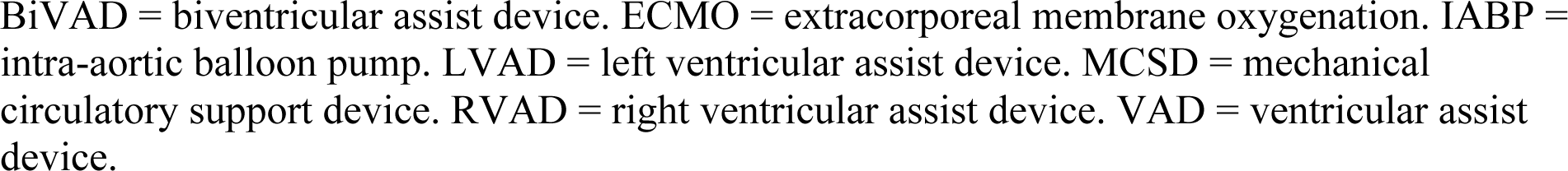
Comparison of Heart Transplant Allocation Before and After Score Change.

Prior to this score change, candidates living with durable LVADs were assigned equal priority to critical patients on veno-arterial extracorporeal membrane oxygenation (VA-ECMO).^3^ LVAD technology, especially with the advent of the magnetically levitated centrifugal-flow HeartMate 3 (HM3), has improved so successfully that a majority of patients are alive after five years with significantly reduced complication rates.^4^ Several studies have described various outcomes since the allocation score change including decreased deterioration of candidates on the waitlist, yet significantly decreased utilization of LVADs and post-transplant survival.^5,6^ However, survival data in particular has been limited thus far given the relative recency of the UNOS policy change. Presently, five-year survival data is available for both patients that received the most advanced and life prolonging LVAD technology and heart transplant recipients after the score change. It is valuable to investigate the impact of policy change on utilization and outcomes of these MCSDs and expand on prior studies showing some alarming trends.

### Objectives

Our primary objective in this study was to compare survival outcomes and implantation strategy between LVAD patients who received a heart transplant before and after the change in allocation policy. We also compared patient demographics, transplant parameters, and LVAD implantation between the groups.

## Methods

### Study design

This is a retrospective review of the UNOS registry to evaluate LVAD utilization and survival outcomes among LVAD patients who received a heart transplant before and after the allocation score change. Data were obtained through the UNOS registry and were entered into an online Health Insurance Portability and Accountability Act-compliant database and de-identified for analysis. The Institutional Review Board at Temple University waived approval for this study since the UNOS dataset contains de-identified information.

### Setting

A total of 4599 patients were identified through the UNOS registry who had an LVAD at the time of listing and/or at the time of transplant. Those transplanted between January 1, 2016 and March 10, 2020 were included. Recipients then were stratified into two groups based on transplantation before or after the allocation score change on October 18, 2018. Those transplanted from January 1, 2016 to October 17, 2018 were in the “before” group; those transplanted from October 18, 2018 to March 10, 2020 were in the “after” group. Survival and follow-up information extended through March 30, 2023.

### Participants

Eligibility criteria included LVAD patients ≥18 years of age who received a heart transplant. Those listed prior to the score change date but transplanted after were excluded. Other exclusion criteria were simultaneous heart-lung transplant, re-transplant recipients, and those missing follow-up or survival information.

### Variables

We collected recipient and donor demographic variables including age, sex, ethnicity, BMI, height, HCV serostatus, cause of death, and CDC risk status. Clinical variables included days on the waitlist, distance from donor hospital to transplant center, length of status, graft status, cause of graft failure, ECMO at listing, LVAD implantation at listing, allocation type, and post-operative complications.

### Data sources/measurement

Baseline recipient characteristics, donor characteristics, and clinical parameters were collected from UNOS. Continuous variables were compared using two-sample t-tests and were reported as mean and standard deviation; categorical variables were compared using chi-squared tests and were reported as counts and percentages.

### Statistical methods

The primary outcome of interest was survival. Survival time was calculated from the date of transplant to the last date of follow-up and was assessed up to 5 years post-transplant. Survival was analyzed with Kaplan-Meier curves and log-rank tests. Secondary outcomes included length of stay, graft status, and post-transplant complications.

A multivariate Cox proportional hazards model was used to identify variables that were significant predictors of mortality. Covariates used were era of transplantation (before versus after the score change), recipient age, donor age, ischemic time, and ECMO status at registration. P-values <0.05 were considered significant. Statistical analyses were conducted with JMP 16.1 (SAS Institute Inc, Cary, NC).

## Results

### Descriptive Data

#### Recipient Demographic Variables

Out of 4599 LVAD patients included in the study, 3767 patients (84.3%) were transplanted before the allocation score change compared to 832 patients (18.6%) after the score change. Table 2 shows the demographics and baseline characteristics for heart transplant recipients and donors stratified by transplantation before versus after the score change. Among heart transplant recipients, those transplanted before the score change were more likely to be older (54.1 versus 53.1 years, p=0.035), male (80.4% versus 75.1%, p=0.0007), and taller in height (175.2 versus 174.3 cm, p=0.0035) compared to those transplanted after the score change.

**Table 2.** Recipient and Donor Demographics and Baseline Characteristics.

| Variable | Before (n=3767) | After (n=832) | P-value |
| --- | --- | --- | --- |
| Recipient age | 54.1 (11.9) | 53.1 (12.2) | 0.0350 |
| Recipient sex |  |  | 0.0007 |
| - Male | 3028 (80.4) | 625 (75.1) |  |
| - Female | 739 (19.6) | 207 (24.9) |  |
| Recipient ethnicity |  |  | 0.9177 |
| - White | 2342 (62.2) | 518 (62.3) |  |
| - Black | 955 (25.4) | 207 (24.9) |  |
| - Asian | 115 (3.1) | 25 (3.0) |  |
| - Hispanic | 313 (8.3) | 75 (9.0) |  |
| - Other/unknown | 42 (1.1) | 7 (0.8) |  |
| Recipient BMI (kg/m<sup>2</sup>) | 28.9 (4.8) | 28.7 (5.1) | 0.2808 |
| Recipient height (cm) | 175.2 (9.6) | 174.1 (9.7) | 0.0035 |
| Recipient HCV serostatus |  |  | 0.3584 |
| - Positive | 115 (3.1) | 24 (2.9) |  |
| - Negative | 3583 (95.1) | 800 (96.2) |  |
| - Not done | 60 (1.6) | 7 (0.8) |  |
| - Unknown | 9 (0.2) | 1 (0.1) |  |
| Donor age | 31.7 (10.5) | 32.7 (10.7) | 0.0086 |
| Donor sex |  |  | 0.0535 |
| - Male | 2868 (76.1) | 607 (73.0) |  |
| - Female | 899 (23.9) | 225 (27.0) |  |
| Donor ethnicity |  |  | 0.1285 |
| - White | 2506 (66.5) | 548 (65.9) |  |
| - Black | 609 (16.2) | 115 (13.8) |  |
| - Asian | 57 (1.5) | 19 (2.3) |  |
| - Hispanic | 553 (14.7) | 138 (16.6) |  |
| - Other/unknown | 42 (1.1) | 12 (1.4) |  |
| Donor BMI (kg/m<sup>2</sup>) | 28.0 (6.0) | 28.4 (6.3) | 0.1024 |
| Donor height (cm) | 175.4 (9.2) | 174.3 (9.0) | 0.0018 |
| Donor cause of death |  |  | <0.0001 |
| - Anoxia | 1363 (36.2) | 384 (46.2) |  |
| - Cerebrovascular/stroke | 525 (13.9) | 113 (13.6) |  |
| - CNS tumor | 17 (0.5) | 1 (0.1) |  |
| - Head trauma | 1791 (47.5) | 307 (36.9) |  |
| - Other/unknown | 71 (1.9) | 27 (3.3) |  |
| Donor CDC risk status |  |  | 0.0002 |
| - Increased risk | 1140 (30.3) | 312 (37.5) |  |
| - Standard risk | 2625 (69.7) | 520 (62.5) |  |
| - Unknown | 2 (0.1) | 0 (0.0) |  |

Continuous variables were reported as mean (standard deviation). Categorical variables were reported as count (percentage). P-values <0.05 were considered significant.

### Donor Demographic Variables

Among heart transplant donors, those before the score change were more likely to be younger (31.7 versus 32.7 years, p=0.0086), taller in height (175.4 versus 174.3 cm, p=0.0018), and standard risk (as opposed to increased risk) CDC status (69.7% versus 62.5%, p=0.0002) compared to those transplanted after the score change. Donor cause of death was significantly different between the two groups (p=0.002). Donors before the score change were less likely to have anoxia (36.2% versus 46.2%) and more likely to have head trauma (47.5% versus 36.9%) as a cause of death compared to donors after the change.

### Recipient Clinical Variables

Table 3 shows the clinical parameters for heart transplant recipients before and after the score change. Recipients before the score change spent more days on the waitlist (369.0 versus 83.5 days, p<0.0001), received organs from shorter distances to the transplant center (143.2 versus 259.5 nautical miles, p<0.0001), and had decreased ischemic time (3.0 versus 3.5 hours, p<0.0001) compared to recipients after the score change. Allocation type was different between the two groups (p<0.0001), with recipients before the score change more likely to have a local allocation compared to recipients after (68.4% versus 35.0%).

**Table 3.** Recipient Clinical Parameters.

| Variable | Before (n=3767) | After (n=832) | P-value |
| --- | --- | --- | --- |
| Days on the waitlist | 369.0 (458.5) | 83.5 (103.5) | <0.0001 |
| Distance from donor hospital to transplant center (nautical miles) | 143.2 (182.1) | 259.5 (246.8) | <0.0001 |
| Length of stay (days) | 23.2 (24.4) | 25.0 (27.4) | 0.0667 |
| Graft status at follow-up |  |  | 0.0331 |
| - Functioning | 3516 (94.0) | 795 (95.9) |  |
| - Failed | 224 (6.0) | 34 (4.1) |  |
| Cause of graft failure |  |  | 0.0485 |
| - Acute rejection | 31 (0.8) | 8 (1.0) |  |
| - Chronic rejection | 45 (1.2) | 3 (0.4) |  |
| - Primary non-functioning | 79 (2.1) | 17 (2.0) |  |
| - Other | 66 (1.8) | 6 (0.7) |  |
| - Unknown or not failed | 3546 (94.1) | 798 (95.9) |  |
| ECMO at registration | 37 (1.0) | 26 (3.1) | <0.0001 |
| ECMO at transplant | 20 (0.5) | 33 (4.0) | <0.0001 |
| Ischemic time (hours) | 3.0 (1.1) | 3.5 (1.1) | <0.0001 |
| LVAD implantation during listing | 1301 (34.5) | 161 (19.4) | <0.0001 |
| Allocation type |  |  | <0.0001 |
| - Local | 2577 (68.4) | 291 (35.0) |  |
| - Regional | 452 (12.0) | 192 (23.1) |  |
| - National | 733 (19.5) | 348 (41.8) |  |
| - Foreign | 5 (0.1) | 1 (0.1) |  |
| Dialysis prior to discharge? |  |  | 0.0386 |
| - Yes | 520 (13.8) | 142 (17.1) |  |
| - No | 3244 (86.1) | 690 (82.9) |  |
| - Unknown | 3 (0.1) | 0 (0.0) |  |
| Permanent pacemaker implanted prior to discharge? |  |  | 0.8591 |
| - Yes | 87 (2.3) | 21 (2.5) |  |
| - No | 3673 (97.5) | 810 (97.4) |  |
| - Unknown | 7 (0.2) | 1 (0.1) |  |
| Stroke prior to discharge? |  |  | <0.0001 |
| - Yes | 134 (3.6) | 59 (7.1) |  |
| - No | 3624 (96.2) | 769 (92.4) |  |
| - Unknown | 9 (0.2) | 4 (0.5) |  |
| LVAD brand at time of transplant |  |  | <0.0001 |
| - HeartMate 2 | 1731 (46.0) | 131 (15.8) |  |
| - HeartMate 3 | 78 (2.1) | 277 (33.3) |  |
| - HeartWare HVAD | 1461 (38.8) | 236 (28.4) |  |
| - Other/unknown | 497 (13.2) | 188 (22.6) |  |

Continuous variables were reported as mean (standard deviation). Categorical variables were reported as count (percentage). P-values <0.05 were considered significant.

### Recipient Post-Operative Complications

Recipients before the score change had more graft failures at follow-up (6.0% versus 4.1%, p=0.0331) and different causes of graft failure compared to recipients after (p=0.0485). Notably, graft failure due to chronic rejection was higher among recipients before versus after the change (1.2% versus 0.4%). Furthermore, recipients before the change had a lower likelihood of post-operative complications including renal failure requiring dialysis (13.8% versus 17.1%, p=0.0386) and stroke (3.6% versus 7.1%, p<0.0001).

### Recipient MCSD Variables

Recipients before the score change were more likely to have LVAD implantation during listing compared to recipients after (34.5% versus 19.4%, p<0.0001). LVAD device types differed between the two groups (p<0.0001). Recipients before the score change had a lower proportion of Heartmate 3 (2.1% versus 33.3%) and higher proportion of Heartmate 2 (46.0% versus 15.8%) and HeartWare HVAD (38.8% versus 28.4%) utilization. ECMO utilization at listing was lower among recipients transplanted before the score change compared to after (0.5% versus 4.0%, p<0.0001).

### Outcomes Data Survival Analysis

There was significantly decreased short-term and long-term survival when comparing patients transplanted before and after the score change. At 90-days, survival in the before group was 94.4%, compared to 91.4% after (p=0.0014). Survival was also significantly increased in the before group compared to the after group at 1-year (91.6% versus 87.6%, p=0.0003), 3-years (85.9 % versus 80.5%, p=0.0002), and 5-years (78.9% versus 60.5%, p<0.0001) following transplant. Figure 1 shows the 5-year Kaplan-Meier survival curve for patients before and after the score change. While survival was significantly different between the groups at all time points, we noted a steeper decline at 3 years for patients transplanted after the score change.

**Figure 1.**
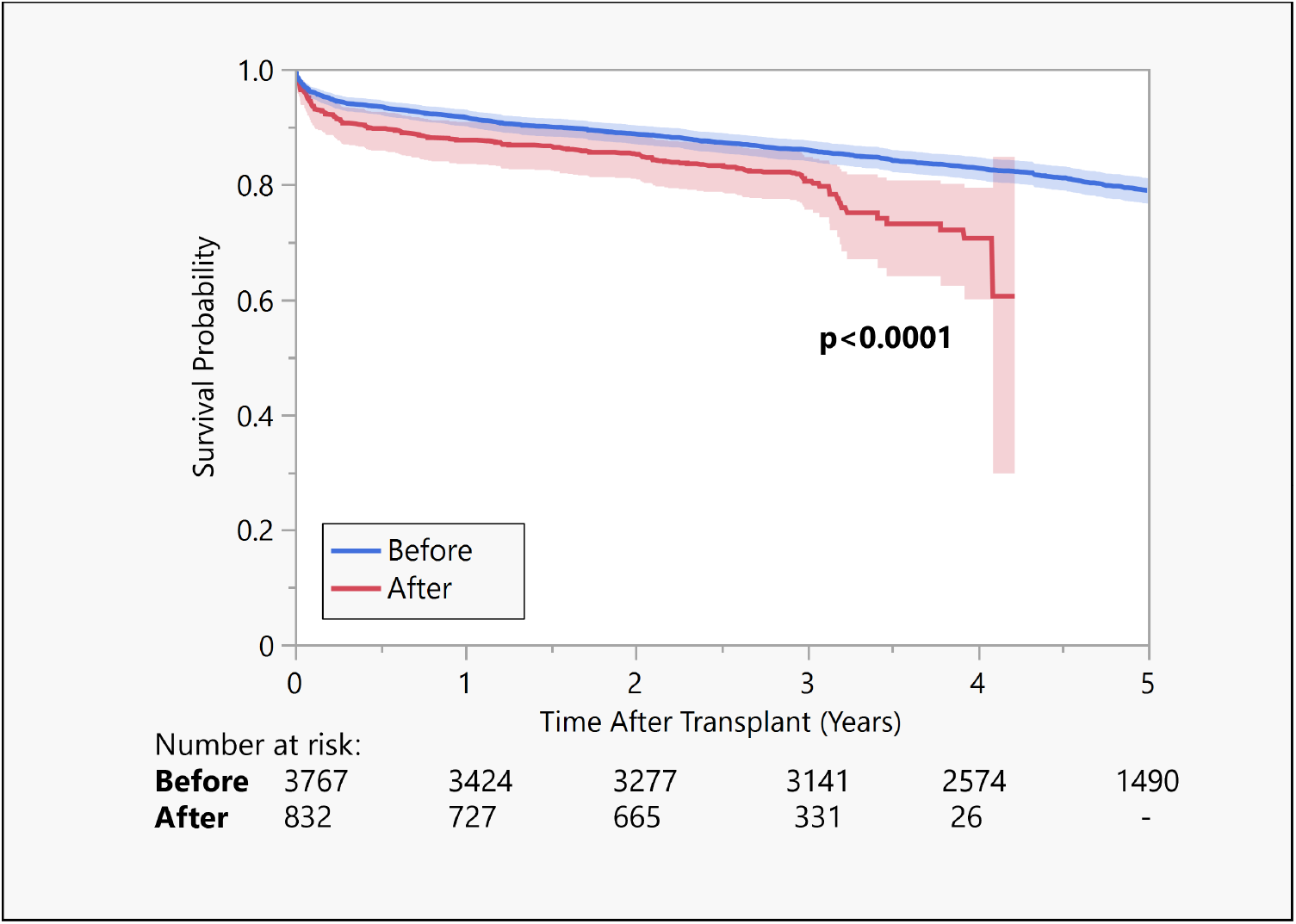
5-year Kaplan-Meier Survival Curve for Before versus After Score Change Compared to LVAD patients transplanted before the 2018 score change, LVAD patients transplanted after demonstrated significantly worsened survival at 5 years post-transplant (p<0.0001).

### Multivariate Cox Proportional Hazards Model

Table 4 shows the multivariate Cox regression model. Compared to transplantation before the score change, transplantation after the score change was associated with increased mortality (HR 1.37, 95% CI 1.14-1.63, p=0.0007). Other predictors of mortality include ischemic time (HR 1.13, 95% CI 1.06-1.19, p<0.0001), ECMO at transplant (HR 1.86, 95% CI 1.12-3.70, p=0.0160), and recipient age (HR 1.01, 95% CI 1.01-1.02, p=0.0002). Donor age was not significantly associated with survival.

**Table 4.** Multivariate Cox proportional hazards model.

| Predictor | Hazard Ratio | 95% Confidence Interval | P-value |
| --- | --- | --- | --- |
| Before score change | 1.00 | Reference | Reference |
| After score change | 1.37 | 1.14-1.63 | 0.0007 |
| Ischemic time | 1.13 | 1.06-1.19 | <0.0001 |
| Recipient age | 1.01 | 1.01-1.02 | 0.0001 |
| Donor age | 1.01 | 1.00-1.02 | 0.0018 |
| No ECMO at transplant | 1.00 | Reference | Reference |
| On ECMO at transplant | 1.86 | 1.12-3.70 | 0.0160 |

## Discussion

We performed a retrospective study of long-term survival outcomes for LVAD patients who received a heart transplant before and after the allocation score change. We demonstrated multiple changes that have occurred since the implementation of this new policy, including decreased short- and long-term survival, increased ischemic time, increased ECMO utilization, and decreased LVAD implantation during listing among patients after the score change compared to before. In converting from a 3-tier to a 6-tier system, the heart transplant allocation score change has created greater stratification based on relative urgency of waitlisted patients, especially given the high volume and heterogeneity of patients categorized as status 1A (most urgent) under the prior policy.^7^ Patients with LVAD as a bridge to heart transplant encompassed a substantial portion of waitlisted patients under the prior policy, in part due to advancements in LVAD technology that have led to their decreased waitlist mortality.^8,9^

For example, the introduction of the HM3 in 2017 has had profound impacts on the management of patients with heart failure. The final report for the MOMENTUM 3 trial published in 2019 demonstrated a less frequent need for pump replacement and lower incidence of complications such as strokes and major bleeding compared to the previous HeartMate 2 (HM2) model.^17^ In the follow-up study to this trial published in 2022, HM3 patients were reported to have a better composite outcome and overall higher likelihood of survival at five years (54.0%) compared to HM2 patients (29.7%).^4^ Ultimately, this led to stable LVAD patients having lower waitlist mortality compared to other status 1A patients under the prior allocation policy and contributed to their deprioritization upon score change. However, given the recency of these advancements in LVAD technology, and the recency of the heart transplant allocation score change, there is limited data on how this change has impacted the long-term survival of LVAD patients.

Our study showed decreased short-term and long-term survival with the new allocation policy. For LVAD patients transplanted prior to the score change, 90-day survival was 94.4% and 5-year survival was 78.9%. For LVAD patients transplanted after the score change, 90-day survival was 91.4% and 5-year survival was 60.5%. Our multivariate analysis demonstrated a statistically significantly increased hazard ratio in those transplanted after the score change (Table 4). These findings build upon the paucity of published long-term survival data among these patients while helping to clarify inconclusive short-term survival data. A study by Liu et al in 2021 compared 38 LVAD patients listed within one year prior to the score change with 33 LVAD patients listed within one year after the score change and found no significant change in post-transplant survival.^3^ A larger study by Jani et al in 2021 compared 1229 LVAD patients listed prior to score change with 955 LVAD patients listed after the score change. They demonstrated comparable 6-month post-transplant survival rates (93.2% and 91.5%, respectively) between the two groups.^8^

On the contrary, a study conducted by Mullan et al in 2021 compared outcomes between 983 patients with LVAD as a bridge to heart transplant in the pre-change period to 814 patients in the post-change period. They demonstrated significantly worse 1-year survival among post-change patients compared to pre-change patients (83.4% versus 91.7%).^12^ Another study by Hess et al in 2023 compared survival outcomes up to two years post-transplant and found no significant differences between the pre-score change (N=1418) and post-score change (N=1142) LVAD patients (90.5% survival versus 90.3% survival, respectively), however they found higher rates of post-transplant stroke and renal failure, as well as a longer hospital length of stay among post-score change patients.^13^ Our findings help clarify the discrepancies described in these prior studies by better characterizing long-term outcomes for LVAD patients. Our study also raises important questions regarding why these patients demonstrated decreased survival under the new allocation policy.

Potential explanations for these survival differences are likely multifactorial. As organs became shared more widely after the allocation policy change, this contributed to increased transport times for donor organs and potential increased risk for ischemia-reperfusion injury and poor outcomes. We found a significant difference in average transport distance between LVAD patients transplanted before (143.2 nautical miles) and after the score change (259.5 nautical miles) and subsequently a difference in average ischemic time between the two groups (3.04 versus 3.47 hours, respectively) (Table 3). Longer ischemic times are associated with higher rates of primary graft dysfunction and mortality after heart transplantation.^14^ While an ischemic time of less than four hours is generally accepted as a threshold for optimal heart transplant outcomes, there is data to suggest that survival may be compromised at lower thresholds.^15^ Likewise, our multivariate analysis demonstrated increased mortality due to longer ischemic time (Table 4).

We also demonstrated differences in ECMO utilization at listing between LVAD patients before and after the score change. Patients before the score change had lower rates of ECMO utilization at transplant (0.5%) compared to patients after (4.0%). This is not entirely unexpected given that patients on ECMO were promoted in status under the new allocation policy. MCSDs such as ECMO have typically been associated with poor outcomes following heart transplantation.^14,16^ However, there is conflicting data on how ECMO is associated with survival in the post-score change era.

A study by Hess et al in 2020 comparing 72 ECMO patients before score change to 93 ECMO patients after score change found no difference in one-year survival rates between the groups (90.3% versus 79.3%, respectively).^17^ Another study by Kim et al in 2022 found that ECMO usage as a bridge to heart transplant was associated with increased mortality compared to non-ECMO patients prior to score change but not after the score change.^18^ These data suggest that increased ECMO utilization may not be the driver of decreased survival among LVAD patients in our post-score change cohort. However, we found that utilization of ECMO at time of transplant is associated with increased mortality in our multivariate analysis (Table 4).

In addition to our survival findings, we demonstrated a decreased rate of LVAD implantation among patients after the score change compared to before, representing a significant shift in the strategies for bridging patients to transplant. Mullan et al suggested that LVAD implantation prior to the score change may have served to increase priority and expedite transplant for certain patients.^12^ LVAD implantation and temporary MCSD utilization strategy has shifted since the allocation score change, and we demonstrate that these trends may contribute to worse outcomes over time.

The improved morbidity and mortality of HM3 lends it useful as a durable bridge to transplant with most candidates transplanted as category 4 or category 3 during the 30-day discretionary period, as described by Uriel et al.^19^ Hawkins et al demonstrated a 1050% increase in use of temporary MSCDs (intra-aortic balloon pump, percutaneous VAD, or VA-ECMO) with concurrent 54% decrease in use of VAD in their population after the allocation score change.^20^ Certain candidates may benefit from ECMO or other temporary MCSDs and the resultant prioritized transplant listing status compared to remaining longer on the waitlist with LVAD alone.

Post-transplant survival after the 2018 score change was reduced among all recipients, not limited to LVAD patients.^5^ One potential contributing variable is the Covid-19 pandemic. However, a 2021 review of Covid-19 outcomes in solid organ transplant recipients demonstrated comparable mortality to the general population.^21^ Further research is warranted to investigate the impact of Covid-19 on outcomes in heart recipients with LVAD.

Although LVAD therapy has significantly improved survival and quality of life for patients with heart failure, heart transplant remains the definitive treatment modality. These findings suggest that the risks and benefits of LVAD implantation and strategy to use them as a bridge to heart transplant may require re-evaluation under the new allocation system, given the significant effect that this policy change has had on post-transplant survival among LVAD patients, in part by promoting riskier bridging strategies.

### Limitations

Despite the thoroughness of our investigation, there may be limitations. This is a retrospective study of LVAD patients from the UNOS registry who received heart transplants before and after the allocation score change. As a multicenter registry, the UNOS registry is susceptible to errors including data entry and missing data. Furthermore, this is a retrospective study susceptible to selection bias as patients listed for transplant were pre-selected by individual institutions as suitable transplant recipients. To control for potential confounders, we conducted a multivariate Cox proportional hazards regression of variables associated with survival, however it is possible that unmeasured factors may have influenced the study’s findings. For example, the UNOS registry does not include data regarding patient adherence or the frequency of LVAD device-related complications between the two groups.

## Conclusion

In this retrospective study of UNOS registry data, we compared LVAD outcomes and strategy among patients transplanted before and after the 2018 heart transplant allocation score change. Our findings of lower 5-year survival and decreased LVAD implantation among patients transplanted after the score change suggest a shift in bridging strategies and raise important concerns regarding the appropriate utilization of LVAD and other MCSDs under the new policy.

## Data Availability

The data that support the findings of this study are available from the United Network for Organ Sharing

## Non-standard Abbreviations and Acronyms

BiVAD: biventricular assist device
BMI: body mass index
CDC: Centers for Disease Control and Prevention
ECMO: extracorporeal membrane oxygenation
HCV: hepatitis C virus
HM2: HeartMate 2
HM3: HeartMate 3
HVAD: HeartWare ventricular assist device
IABP: intra-aortic balloon pump
LVAD: left ventricular assist device
MCSD: mechanical circulatory support device
RVAD: right ventricular assist device
UNOS: United Network for Organ Sharing
VAD: ventricular assist device
VA-ECMO: veno-arterial extracorporeal membrane oxygenation

## Acknowledgements

None.

## References

1. Tran Z, Hernandez R, Madrigal J, Kim ST, Verma A, Rabkin DG, Benharash P. Center-Level Variation in Transplant Rates Following the Heart Allocation Policy Change. JAMA Cardiol. 2022 Mar 1;7(3):277–285. doi: 10.1001/jamacardio.2021.5370. PMID: 35044415; PMCID: PMC8771429.

2. Liz Robbins C. OPTN/UNOS proposal to modify the adult heart allocation system. 2016 Available at https://optn.transplant.hrsa.gov.pdf.

3. Liu J, Yang BQ, Itoh A, Masood MF, Hartupee JC, Schilling JD. Impact of New UNOS Allocation Criteria on Heart Transplant Practices and Outcomes. Transplant Direct. 2020 Dec 15;7(1):e642. doi: 10.1097/TXD.0000000000001088. PMID: 33335981; PMCID: PMC7738116.

4. Mehra MR, Goldstein DJ, Cleveland JC, Cowger JA, Hall S, Salerno CT, Naka Y, Horstmanshof D, Chuang J, Wang A, Uriel N. Five-Year Outcomes in Patients With Fully Magnetically Levitated vs Axial-Flow Left Ventricular Assist Devices in the MOMENTUM 3 Randomized Trial. JAMA. 2022 Sep 27;328(12):1233–1242. doi: 10.1001/jama.2022.16197. PMID: 36074476; PMCID: PMC9459909.

5. Kilic A, Mathier MA, Hickey GW, Sultan I, Morell VO, Mulukutla SR, Keebler ME. Evolving Trends in Adult Heart Transplant With the 2018 Heart Allocation Policy Change. JAMA Cardiol. 2021 Feb 1;6(2):159–167. doi: 10.1001/jamacardio.2020.4909. PMID: 33112391; PMCID: PMC7593876.

6. Yuzefpolskaya M, Schroeder SE, Houston BA, Robinson MR, Gosev I, Reyentovich A, Koehl D, Cantor R, Jorde UP, Kirklin JK, Pagani FD, D’Alessandro DA. The Society of Thoracic Surgeons Intermacs 2022 Annual Report: Focus on the 2018 Heart Transplant Allocation System. Ann Thorac Surg. 2023 Feb;115(2):311–327. doi: 10.1016/j.athoracsur.2022.11.023. Epub 2022 Dec 1. PMID: 36462544.

7. Dardas T, Mokadam NA, Pagani F, Aaronson K, Levy WC. Transplant registrants with implanted left ventricular assist devices have insufficient risk to justify elective organ procurement and transplantation network status 1A time. J Am Coll Cardiol. 2012 Jul 3;60(1):36–43. doi: 10.1016/j.jacc.2012.02.031. Epub 2012 Apr 25. PMID: 22541833.

8. Schulze PC, Kitada S, Clerkin K, Jin Z, Mancini DM. Regional differences in recipient waitlist time and pre- and post-transplant mortality after the 2006 United Network for Organ Sharing policy changes in the donor heart allocation algorithm. JACC Heart Fail. 2014 Apr;2(2):166–77. doi: 10.1016/j.jchf.2013.11.005. PMID: 24720925; PMCID: PMC4283198.

9. Pinney SP. Timing isn’t everything: donor heart allocation in the present LVAD era. J Am Coll Cardiol. 2012 Jul 3;60(1):52–3. doi: 10.1016/j.jacc.2012.03.017. Epub 2012 Apr 25. PMID: 22541832.

10. Mehra MR, Uriel N, Naka Y, Cleveland JC Jr, Yuzefpolskaya M, Salerno CT, Walsh MN, Milano CA, Patel CB, Hutchins SW, Ransom J, Ewald GA, Itoh A, Raval NY, Silvestry SC, Cogswell R, John R, Bhimaraj A, Bruckner BA, Lowes BD, Um JY, Jeevanandam V, Sayer G, Mangi AA, Molina EJ, Sheikh F, Aaronson K, Pagani FD, Cotts WG, Tatooles AJ, Babu A, Chomsky D, Katz JN, Tessmann PB, Dean D, Krishnamoorthy A, Chuang J, Topuria I, Sood P, Goldstein DJ; MOMENTUM 3 Investigators. A Fully Magnetically Levitated Left Ventricular Assist Device - Final Report. N Engl J Med. 2019 Apr 25;380(17):1618–1627. doi: 10.1056/NEJMoa1900486. Epub 2019 Mar 17. PMID: 30883052.

11. Jani M, Lee S, Hoeksema S, Archarya D, Boeve T, Manandhar-Shrestha N, Leacche M, Jovinge S, Loyaga-Rendon R. Changes in wait list mortality, transplantation rates and early post-transplant outcomes in LVAD BTT with New Heart Transplant Allocation Score. A UNOS database analysis. The Journal of Heart and Lung Transplantation. 2021 Apr 21;40(4):S17 doi: 10.1016/j.healun.2021.01.1776

12. Mullan CW, Chouairi F, Sen S, Mori M, Clark KAA, Reinhardt SW, Miller PE, Fuery MA, Jacoby D, Maulion C, Anwer M, Geirsson A, Mulligan D, Formica R, Rogers JG, Desai NR, Ahmad T. Changes in Use of Left Ventricular Assist Devices as Bridge to Transplantation With New Heart Allocation Policy. JACC Heart Fail. 2021 Jun;9(6):420–429. doi: 10.1016/j.jchf.2021.01.010. Epub 2021 Mar 10. PMID: 33714748.

13. Hess NR, Ziegler LA, Keebler ME, Hickey GW, Kaczorowski DJ. Impact of 2018 allocation system change on outcomes in patients with durable left ventricular assist device as bridge to transplantation: A UNOS registry analysis. J Heart Lung Transplant. 2023 Jul;42(7):925–935. doi: 10.1016/j.healun.2023.02.002. Epub 2023 Feb 10. PMID: 36973093.

14. Russo MJ, Iribarne A, Hong KN, Ramlawi B, Chen JM, Takayama H, Mancini DM, Naka Y. Factors associated with primary graft failure after heart transplantation. Transplantation. 2010 Aug 27;90(4):444–50. doi: 10.1097/TP.0b013e3181e6f1eb. PMID: 20622755.

15. Tang PC, Wu X, Zhang M, Likosky D, Haft JW, Lei I, Abou El Ela A, Si MS, Aaronson KD, Pagani FD. Determining optimal donor heart ischemic times in adult cardiac transplantation. J Card Surg. 2022 Jul;37(7):2042–2050. doi: 10.1111/jocs.16558. Epub 2022 Apr 30. PMID: 35488767; PMCID: PMC9325483.

16. Fukuhara S, Takeda K, Kurlansky PA, Naka Y, Takayama H. Extracorporeal membrane oxygenation as a direct bridge to heart transplantation in adults. J Thorac Cardiovasc Surg. 2018 Apr;155(4):1607-1618.e6. doi: 10.1016/j.jtcvs.2017.10.152. Epub 2017 Dec 21. PMID: 29361299.

17. Hess NR, Hickey GW, Sultan I, Kilic A. Extracorporeal membrane oxygenation bridge to heart transplant: Trends following the allocation change. J Card Surg. 2020 Oct 14. doi: 10.1111/jocs.15118. Epub ahead of print. PMID: 33090585.

18. Kim ST, Xia Y, Tran Z, Hadaya J, Dobaria V, Choi CW, Benharash P. Outcomes of extracorporeal membrane oxygenation following the 2018 adult heart allocation policy. PLoS One. 2022 May 20;17(5):e0268771. doi: 10.1371/journal.pone.0268771. PMID: 35594315; PMCID: PMC9122227.

19. Uriel MH, Clerkin KJ, Takeda K, Naka Y, Sayer GT, Uriel N, Topkara VK. Bridging to transplant with HeartMate 3 left ventricular assist devices in the new heart organ allocation system: An individualized approach. J Heart Lung Transplant. 2023 Jan;42(1):124–133. doi: 10.1016/j.healun.2022.08.022. Epub 2022 Sep 10. PMID: 36272893.

20. Hawkins RB, Scott E, Mehaffey JH, Strobel RJ, Speir A, Quader M, Teman NR, Yarboro LT. Influence of heart transplant allocation changes on hospital resource utilization. JTCVS Open. 2022 Nov 4;13:218–231. doi: 10.1016/j.xjon.2022.11.002. PMID: 37063148; PMCID: PMC10091209.

21. Heldman MR, Kates OS. COVID-19 in Solid Organ Transplant Recipients: a Review of the Current Literature. Curr Treat Options Infect Dis. 2021;13(3):67–82. doi: 10.1007/s40506-021-00249-6. Epub 2021 Jun 29. PMID: 34220357; PMCID: PMC8238515.

